# Contributions of age and clinical depression to metacognitive performance

**DOI:** 10.1101/2022.01.27.22269959

**Authors:** Catherine Culot, Tina Lauwers, Carole Fantini-Hauwel, Yamina Madani, Didier Schrijvers, Manuel Morrens, Wim Gevers

## Abstract

**Background:** Pivotal for adaptive behaviour is the ability to judge whether our performance is correct or not, even in the absence of external feedback. This metacognitive ability can be measured using confidence ratings. Past research suggests that overall confidence (confidence bias) is lower in Major Depressive Disorder (MDD). Less is known about the ability of MDD patients to discriminate correct from incorrect performance (metacognitive efficiency).

**Methods:** The perceptual metacognitive performance of 17 aged MDD patients (62-89 years) was tested and compared to an age matched as well as a young control group (21-28 years). The younger control group was included to disentangle the effects of age and MDD on metacognitive performance. Indeed, earlier studies found an increased confidence bias but a decreased metacognitive efficiency with age.

**Results:** Our results showed no difference in confidence bias nor in metacognitive efficiency between MDD patients and age-matched control participants. Importantly, aged participants, including both aged controls and MDD patients, demonstrated lower metacognitive efficiency compared to young participants. This metacognitive deficit found in aged participants was specifically driven by an overconfidence on incorrect trials.

**Conclusions:** The relation between MDD and confidence bias may not be as robust as previously claimed. Furthermore, our results point to the importance of age rather than MDD for metacognitive efficiency. This could have important clinical implications as the observation of lower metacognitive abilities in patients is not necessarily caused by the depression.

**Limitations:** Sample size was relatively small. Some clinical variables such as anxiety, or medication were not controlled.

## 1. Introduction

Major Depressive Disorder (MDD) is a highly prevalent mental disorder that affects more than 320 millions of individuals, or 4.4% of the worldwide population (World Health Organization, 2017). MDD is mainly characterized by a sad mood, apathy, helplessness and ruminations (Kanter et al., 2008; Otte et al., 2016). In the past decade, there has been a surge of interest towards a better understanding of the cognitive factors involved in MDD. Evidence accumulates that impaired metacognition, the ability to reflect on and evaluate one’s own behaviour, is one of these factors (Hoven et al., 2019; Rouault, Seow, et al., 2018; Trauelsen et al., 2016). Some theories, such as the Self-Regulatory Executive Function (S-REF, Wells & Matthews, 1994) argue that dysfunctional metacognitive processes play a causal role in the development and maintenance of mental disorders. According to the S-REF theory, some forms of metacognitive representations could predispose depressive individuals to develop counter-productive response patterns to thoughts and internal events, leading to maladaptive coping strategies (Wells, 2011). One specific response pattern, called the Cognitive-Attention Syndrome (CAS) involves an excessive engagement of worry and rumination, excessive attention to threat and negative information and overly self-focus. It also involves dysfunctional regulation strategies such as avoidance or thought suppression (Sun et al., 2017). The CAS is progressively formed by metacognitive beliefs such as that worries and ruminations are useful, or that thoughts and feelings are something negative and uncontrollable and need to be avoided and suppressed (Sun et al., 2017). The CAS progressively traps the depressive individual in a prolonged state of emotional disturbance that gradually leads to perseverative thinking and contributes to the development and maintenance of the clinical disorder.

The biased metacognition found in depressive individuals does not only concern their thought processes, but also how they evaluate their own performance. Evidence accumulates suggesting that such metacognitive monitoring of performance is impaired in mental disorders and particularly in MDD (Hoven et al., 2019; Rouault, Seow et al., 2018). More specifically, inaccurate confidence about one’s own performance, leading to biased evaluations and detrimental decision-making, are thought to play a role in the development and maintenance of several psychiatric symptoms (Elliott et al., 1996; Hoven et al., 2019; Moritz et al., 2014; Rouault et al., 2018). Using confidence ratings, metacognitive monitoring abilities can be measured by two indices (Fleming and Daw, 2017; Fleming and Lau, 2014; Maniscalco and Lau, 2012). The *confidence bias* (i.e. mean confidence level) reflects the tendency for an individual to report an overall high or low level of confidence. This measure is independent of *metacognitive efficiency*, the ability to discriminate correct from incorrect performance (Fleming and Lau, 2014). For instance, a person can be highly confident in his/her performance (i.e. a high confidence bias) but unable to recognize errors in his/her performance (i.e. low metacognitive efficiency). Non-clinical and clinical studies have shown that confidence bias and metacognitive efficiency can be independently affected by different psychopathological symptoms (Culot et al., 2021a; Culot et al., 2021b; Hoven et al., 2019; Rouault et al., 2018).

Most studies that investigated metacognitive monitoring in depressive patients found a general decrease in confidence bias compared to healthy participants (see Hoven et al., 2019 for a review). This underestimation tendency was demonstrated across different tasks such as memory tasks, general knowledge tasks, perceptual discrimination tasks or social judgement tasks (Fu et al., 2005; Hancock et al., 1996). The confidence bias also seems to correlate with depression severity (Hancock et al., 1996) and improves in recovered patients (Fu et al., 2005). The relation between metacognitive monitoring abilities and depressive symptoms was also investigated in the general population in two recent studies (Moses-Payne et al., 2019; Rouault et al., 2018). Rouault and colleagues (2018) performed a factor analysis to explore the relation between self-reported psychiatric symptoms and metacognitive performance in a general population. High scores on a “depressive/anxious” dimension related to lower confidence biases but higher metacognitive efficiency in a perceptual task. In contrast, high scores on a “compulsive behaviour and intrusive thought” dimension related to the reverse pattern: overconfidence bias but low metacognitive efficiency. Moses-Payne and colleagues (2019) aimed to extend these findings by exploring specifically the association depressive symptoms, self-esteem and metacognitive performance. They observed marginal to no correlation between confidence bias or metacognitive efficiency and depressive scores. Conversely, self-esteem was strongly associated to differences in overall confidence.

Another factor that seems to influence confidence bias and metacognitive efficiency is age. Indeed, several studies suggest an influence of normal aging on cognitive and metacognitive abilities (Palmer et al., 2014; Siegel and Castel, 2019). Healthy older adults have been shown to be overconfident in their performance compared to young participants (Dodson et al., 2007). Perceptual metacognitive efficiency, on the other hand, seems to decline with age, even when task performance is equated across all participants (Palmer et al., 2014). When considering the influence of clinical depression on confidence bias and metacognitive efficiency, age seems to be an important factor that needs to be considered.

Taken together, most studies tend to suggest a reduced confidence bias with depression, even though this finding seems to be more robust in clinical studies than in the general population. Metacognitive efficiency, on the other hand, did not seem to be affected by depression in non-clinical studies. Up until now, metacognitive efficiency was not investigated in clinical populations of MDD patients.

To investigate this question, we compared the confidence bias and metacognitive efficiency of MDD patients with that of control participants. Note that our clinical sample was composed essentially of older MDD patients^1^. Because normal aging has been shown to also impair metacognition, we decided to include two different control groups. This allows to disentangle the respective contributions of age and depression on metacognitive abilities. One control group consisted of older healthy participants paired in age, gender and education level with the MDD patients. The second control group, on the other hand, consisted of young healthy individuals only.

## 2. Material and Method

### 2.1. Participants

Twenty patients (mean age: 75,21 years ± 8.11, 14 women) were recruited at the University Psychiatric Centre Duffel in Belgium. All patients were clinically diagnosed with Major Depressive Disorder by a psychiatrist according to the Diagnostic and Statistical Manual of Mental Disorders V (5^h^ edition, American Psychiatric Association, 2013). Exclusion criteria were (i) comorbidity with other diagnosed psychiatric disorders, drug addiction or anxiety disorder, (ii) history of seizure disorder, brain injury or known neurological disease, (iii) known cognitive disorders, (iii) receiving electroconvulsive therapy 12 months before the study. Three patients were excluded from the analyses (2 clearly misunderstood the task and 1 because of technical problems). The final sample of patients consisted of seventeen patients (mean age: 74.88 years ± 8.11, 12 females). Before starting the experiment, patients completed the Beck Depression Inventory-II (BDI, Beck et al., 1961). The BDI is a 21-items self-report scale measuring the severity of depression with a clinical depression cut-off of 17.

Twenty age- and education-matched healthy control participants (mean age: 75 years ± 7.48, 8 females) were recruited through advertisement. A third group consisting of twenty young healthy control participants (mean age: 23.47 years ± 1.71, 15 females) was recruited from undergraduate psychology course. Control participants were not included if they presented previous psychiatric or substance abuse diagnosis, were taking antidepressant and/or antipsychotic medication or had cognitive or neurological history. Both aged and young control participants completed the BDI-II before the start of the experiment. Participants with a score of 17 or above (cut-off for clinical depression) were excluded from the study (3 aged participants). Aged participants also completed the Montreal Cognitive Assessment (MoCA, Nasreddine et al., 2005), a widely used screening tool for mild cognitive impairments. Participants with a score of below 26 were excluded from the study (1 aged participant). Samples demographics are presented in Table 1. All participants signed informed consent before starting the experiment. The study was approved by the Ethics Committee of the University Hospital of Antwerp on October 19^th^ 2017 (EC1745).

**Table 1.** Demographic and questionnaires for each group (young controls, aged controls and MDD patients). BDI, Beck Depression Inventory.

| | Young Controls<br>(Mean ± SD) | Aged Controls<br>(Mean ± SD) | MDD Patients<br>(Mean ± SD) |
| --- | --- | --- | --- |
| Age | 23.5 ± 1.7 | 74.9 ± 7.25 | 74.7 ± 7.9 |
| Gender (f/m) | 15/5 | 8/8 | 12/5 |
| Education (y) | 12 ± 0 | 12.3 ± 3.2 | 12.01 ± 3.4 |
| BDI | 4.1 ± 3.37 | 5.3 ± 3.9 | 32.1 ± 13.1 |

### 2.2. Metacognitive Task

The task^2^ (Fleming et al., 2014) was programmed with the Psychtoolbox (Brainard, 1997) on MATLAB (MathWorks Inc., Natick, MA, USA) using a modified version of an existing code (Fleming et al., 2014). On each trial, two circles were presented on the screen (diameter 5.1°, eccentricities 8.9°). After 1 second, a variable number of dots (diameter 0.4°) appeared in the circles. The dots disappeared after 1 second. Participants were asked to indicate which one of the two circles contained more dots. Responses were given by thumb presses on handheld buttons connected to a response box (The Black Box Toolkit, Ltd, England) or by telling the experimenter. The difference in number of dots between the two circles was constantly adjusted using a staircase procedure so that all participants had a 70% accuracy rate. Once a response was given, participants were asked to indicate how confident they were in their perceptual judgement. To do so, participants could move a cursor on a continuous sliding scale ranging from 1 to 6. They confirmed their position on the scale by pressing the “space” bar before starting the next trial or by telling the experimenter. The task started with a practice block without the confidence rating in order to familiarize participants and to calibrate the difficulty with the staircase procedure. The task comprised four blocks of 25 trials.

### 2.3. Statistical Analysis

Demographic data of patients and controls were compared using Mann-Whitney U-test and χ^**2**^ -test. Then, the differences in task performance between the three groups were investigated using between-subject ANOVAs and post-hoc pairwise tests with Bonferroni correction. The accuracy was used as a within-subject factor when appropriate. As dependent measures of first-order task-performance, we used the percentage of correct responses, the reaction-times and the “contrast” (average numerical difference between the two circles, reflecting to which difficulty level participants were able to perform at 70% correct). Metacognitive performance was measured by the confidence bias and the metacognitive efficiency (Fleming and Lau, 2014; Maniscalco and Lau, 2012). The confidence bias was simply calculated as the average confidence ratings and reflects the general tendency to be high or low confident. Metacognitive efficiency was calculated using the M-ratio, a common measure of metacognition developed by Maniscalco and Lau (2012) based on the signal detection theory (Green and Swets, 1988). The M-ratio is calculated as the ratio between the metacognitive sensitivity (meta-d’) and the objective, perceptual, sensitivity (d’). This measure is particularly useful as it controls for variation in first-order sensitivity (Fleming & Lau, 2014). To estimate M-ratio (meta-d’/d’) at the group level, we used a recent hierarchical Bayesian modelling approach implemented in a free toolbox (http://www.columbia.edu/~bsm2105/type2sdt/; Fleming, 2017). This toolbox extends the metacognitive model of Maniscalco & Lau (2012) to a Bayesian framework and allows comparison of both subject- and group-level parameters (Fleming, 2017). By incorporating within and between-subject uncertainty, Bayesian estimation is particularly suited for studies with restricted number of trials as in our case (Fleming, 2017; Kruschke, 2014). We estimated metacognitive efficiency for each group separately and then compared their posterior group distributions. Significance of the group-level comparison was assessed by computing the difference of the group posteriors and comparing whether the resulting distribution overlapped with 0. Despite the relatively small number of trials, we also estimated the M-ratio at the subject-level using the non-hierarchical Bayesian model to perform correlational analyses.

These analyses were complemented by a regression linear approach. Finally, Bayesian analyses (Eidswick, 2012) were conducted in case of non-significant results to confirm the absence of effect (null hypothesis).

## 3. Results

### 3.1. Demographic data

No significant differences were found in age, gender or education when comparing the patients to the aged control participants. Young and aged control groups did not differ neither in gender and education level. Contrasts showed, as expected, that patients have significantly higher BDI scores than aged and young control participants (t(50) = 11.77, p < .001). Young and aged control participants did not differ in their BDI scores (t(50) = 0.479, p = .634). Demographic and psychiatric information are depicted in Table 1.

### 3.2. Task performance

The three groups did not differ in task performance (% correct) (*F*(2,50) = 0.352, *p* = .705), demonstrating the efficiency of the staircase procedure setting performance around 70%. The ANOVA on “contrast” (level of difficulty to perform at 70% correct) did not show any significant difference either (*F*(2,50) = 1.593, *p* = .213). Reaction-Times (RTs) differed significantly across groups (*F(*2,50) = 31.73, *p* < .001, *η*_*p*_^*2*^ = 0.56). Post-hoc tests with Bonferroni correction revealed that young participants were faster (1.3 ± 0.7 sec) to perform the perceptual task than the aged control participants (2.4 ± 1.2 sec, *p* < .001) and the MDD patients (2.7 ± 1.1 sec, *p* < .001). Aged control participants did not differ from MDD patients (*p* = .10).

### 3.3. Metacognitive performance

#### Confidence bias

We found a significant effect of group on confidence bias (*F*(2,50) = 4.474, *p* = .0163, *η*_*p*_^*2*^ = 0.15) (Figure 1) indicating that aged control participants reported significantly higher overall confidence levels than young control participants (*p* = .024). MDD patients also reported marginally higher confidence levels than young participants (*p* = .084). Aged control participants and MDD patients did not differ in their confidence bias (*p* = 1.00). Bayesian independent samples t-tests were conducted to further confirm these results on confidence bias. The Bayes-factor (BF) for the comparison between aged and young controls was 8.04 while the BF for the comparison between MDD patients and young control participants was 6.08, providing substantial evidence for the presence of a lower confidence bias in young control participants. When comparing aged controls and MDD patients, a BF of 0.33 was obtained, which does not provide evidence for the presence nor for a difference between these groups.

**Figure 1.**
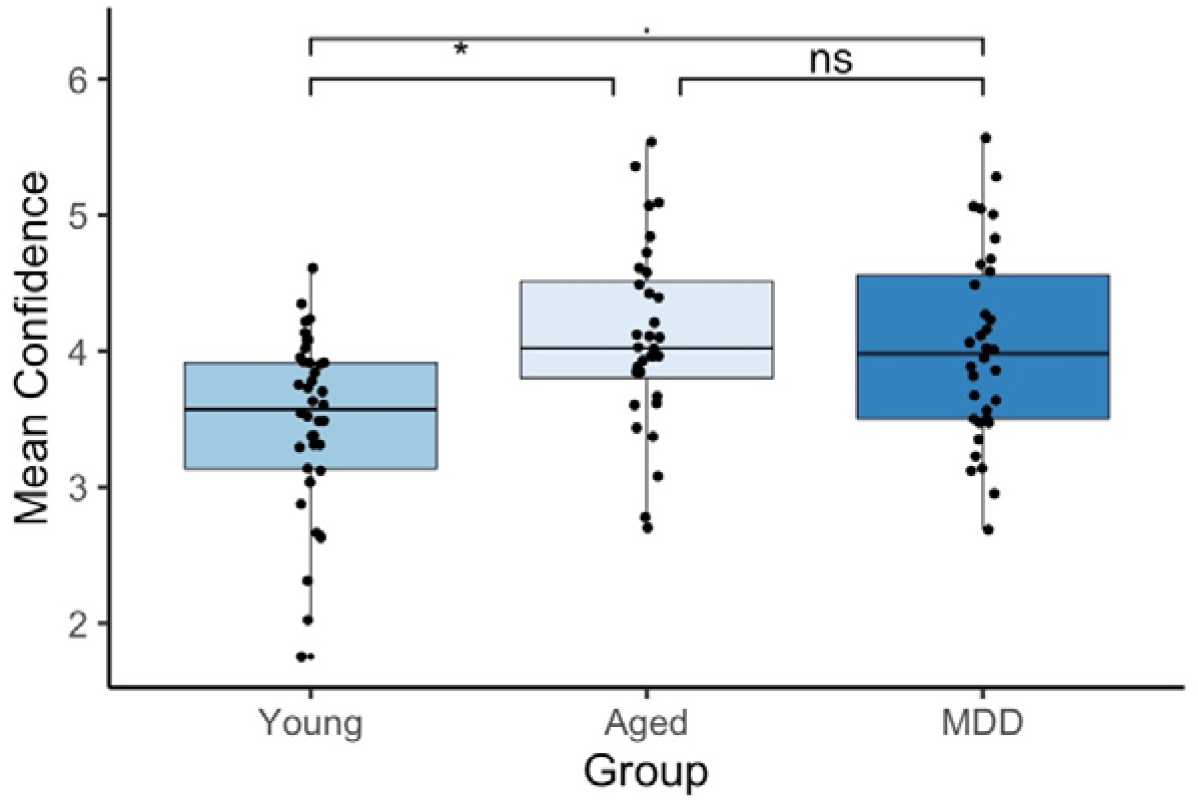
Average confidence levels across groups (Young controls, Aged controls and MDD patients). Confidence was significantly lower for young control participants compared to aged control participants and marginally lower compared to MDD patients. Medium segments indicate median. Upper and lower segments of the boxes indicate third and first quartiles. Whiskers indicate the lowest (highest) values still within the 1.5 interquartile range from the box. * p < .05,. p < .09, ns non-significant.

#### Metacognitive efficiency

Hierarchical Bayesian estimation of group-level metacognitive efficiency revealed that the mean M-ratio was 0.83 for the young controls, 0.61 for the aged controls and 0.48 for the MDD patients. This means that young participants exploited about 80% of the perceptual signal for their metacognitive judgement, aged participants exploited 60% and MDD patients used only about 50% of the perceptual signal. A group comparison (Figure 2A) showed that metacognitive efficiency was significantly lower in MDD patients than in young participants as revealed by the 95% CI of the difference distribution being greater than zero (95% CI: 0.1506 – 0.9886). In contrast, metacognitive efficiency of aged control participants did not differ from the one of the MDD patients (95% CI: -0.268 – 0.762). Young and aged control participants did not differ either (95% CI: -0.0123 – 0.6624).

**Figure 2.**
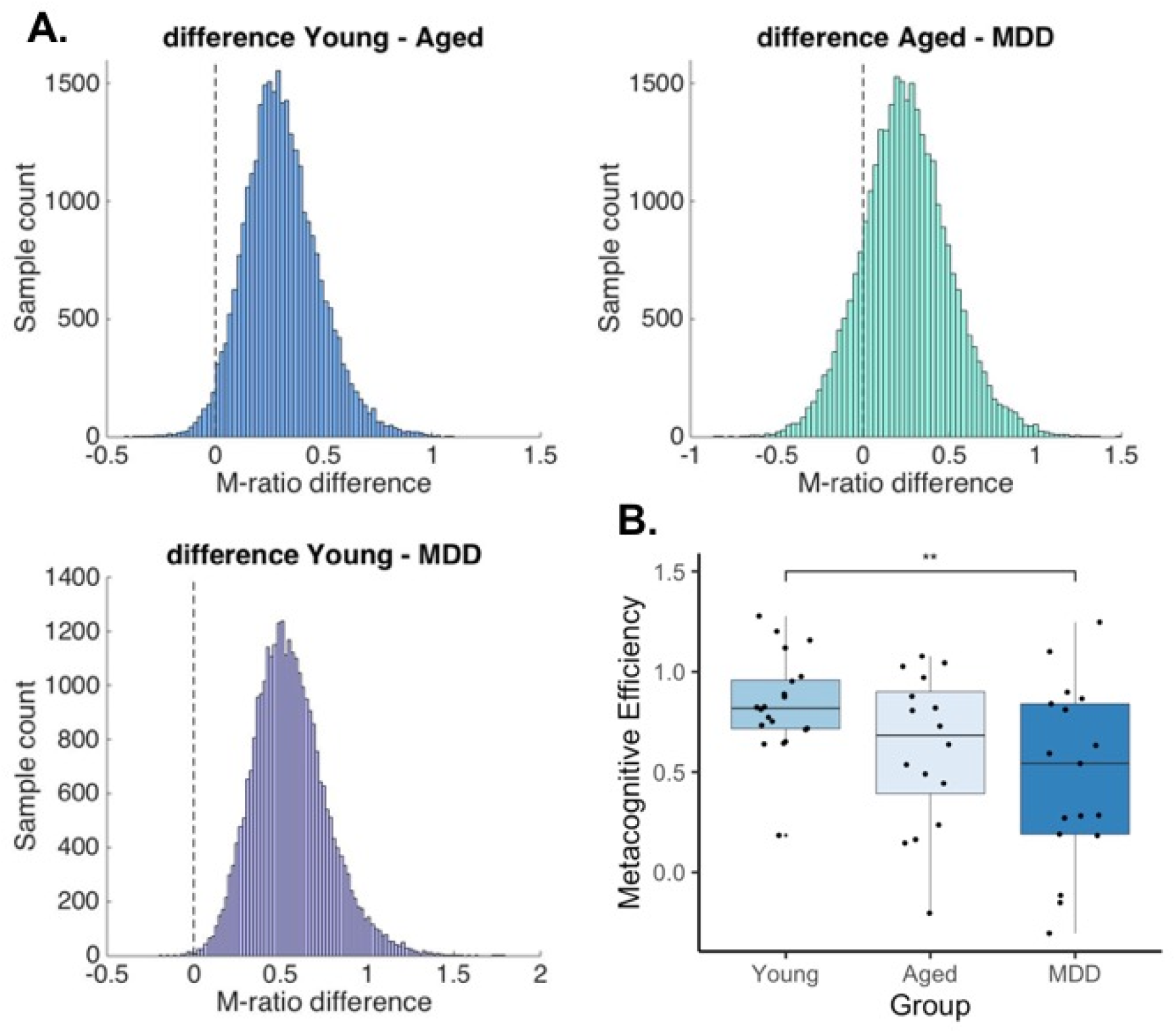
**A**. Difference in group posteriors of metacognitive efficiency estimated by a group-level Bayesian hierarchical model (Fleming, 2017). The difference of distributions between Young controls and MDD patients is significantly different from 0. **B**. Metacognitive efficiency (meta-d’/d’) across the three groups (Young controls, Aged controls, MDD patients). Metacognitive efficiency was lower for MDD patients than for Young controls. Medium segments indicate median. Upper and lower segments of the boxes indicate third and first quartiles. Whiskers indicate the lowest (highest) values still within the 1.5 interquartile range from the box. ** p < .01

These findings were further confirmed by the analysis of the subject-level M-ratio (Figure 2B, note that the calculation of M-ratio is calculated on the basis of Bayesian modelisation) showing a significantly lower metacognitive efficiency for MDD patients than for young participants (*p* = .013) (*F*(2,50) = 4.597, *p* = .015, *η*_*p*_^*2*^ = 0.16).

#### Confidence on correct and error trials

The previous analyses showed that MDD patients had lower metacognitive efficiency compared to young participants. This indicates that MDD patients have more difficulty to discriminate their correct responses from their errors when reporting their confidence. However, the analysis of the M-ratio does not inform about what causes this deficit. Either MDD patients reported lower confidence on their correct trials or they were overconfident on their errors, or both at the same time. To answer this question, we compared the overall confidence across groups looking at both correct and incorrect responses (Figure 3). A significant effect of accuracy (*F*(1,50) = 117.94, *p* < .001), *η*_*p*_^*2*^ = 0.69) was observed indicating that confidence was higher for correct (4.1 ± 1.2) than for incorrect (3.6 ± 1.18) responses. The main effect of group was again significant (*F*(2,50) = 5.74, *p* = .006, *η*_*p*_^*2*^ = 0.24), showing that young participants were overall less confident (3.63 ± 1.3) than aged control (4.2 ± 1.03, *p* < .001) and MDD patients (4.16 ± 1.2, *p* = .002). There was no difference in confidence between aged controls and MDD patients. The significant interaction between group and accuracy (*F*(2,50) = 8.7, *p* < .001, *η*_*p*_^*2*^ = 0.18) revealed that MDD patients and older healthy participants were more confident than young participants, but only when the responses were incorrect (young – aged : p = .006; young – MDD patients: p = .007). No differences between the three groups (all *p* > .400) were observed when the responses were correct. The Bayesian comparison of confidence in correct trials between the three groups resulted in a BF of 1.00, thus not providing evidence for a difference.

**Figure 3.**
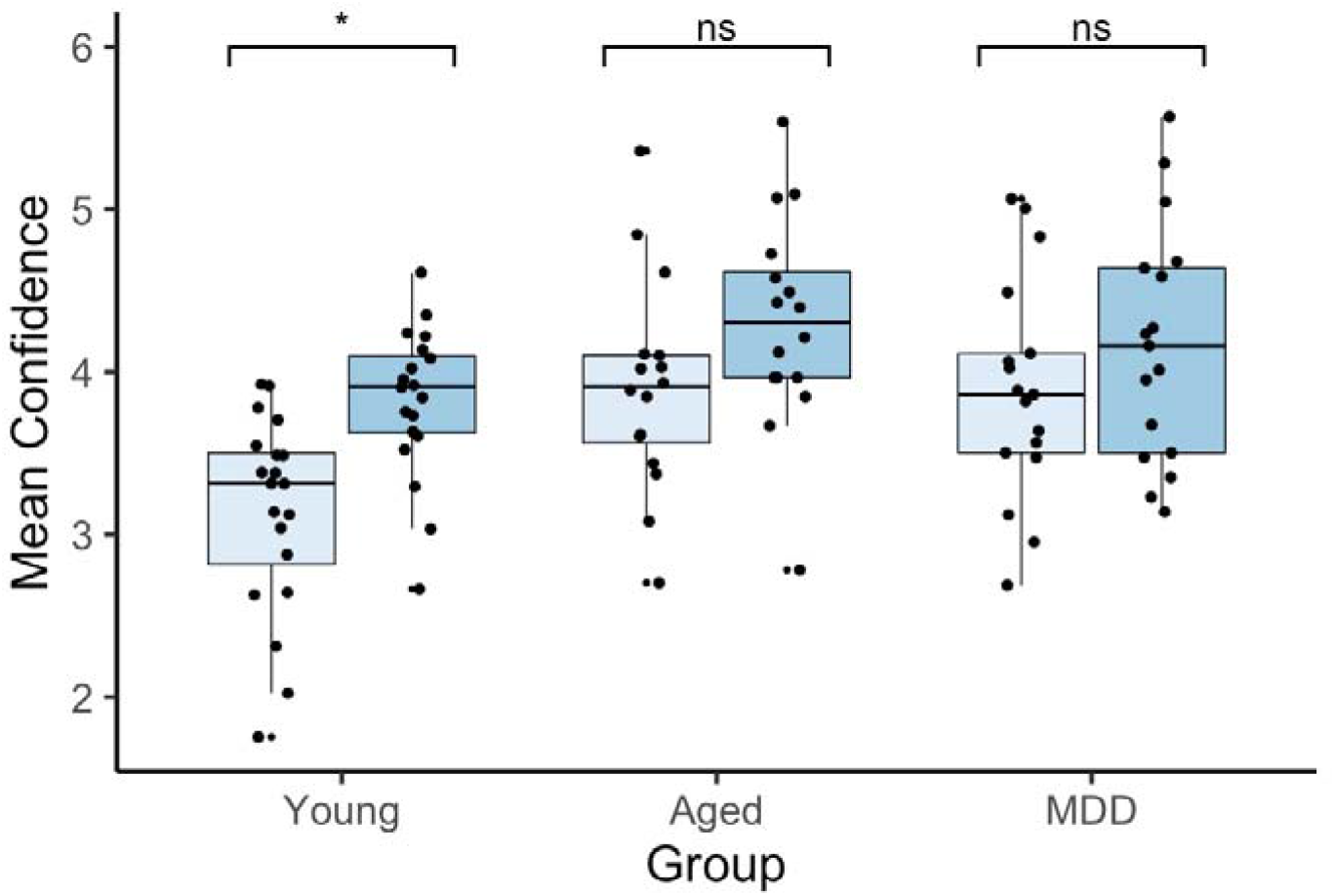
Average confidence levels across groups (Young controls, Aged controls and MDD patients) and accuracy (correct vs incorrect responses). Confidence was lower for incorrect responses only in the young group. Medium segments indicate median. Upper and lower segments of the boxes indicate third and first quartiles. Whiskers indicate the lowest (highest) values still within the 1.5 interquartile range from the box. * p < .05. ns, non-significant.

Finally, only the young participants reported less confidence on incorrect compared to correct responses (correct: 3.8 ± 1.2 – incorrect: 3.1 ± 1.2, p = .011). These results thus suggest that the deficit in metacognitive efficiency found in MDD patients is due to an overconfidence in their incorrect responses rather than to an under-confidence in their correct responses. This overconfidence on incorrect responses was also observed in aged control participants, suggesting that they also have altered metacognitive efficiency, even if it did not reach significance in the analysis of the M-ratio. Note that this overconfidence in errors also explains the higher overall confidence bias found in aged controls and MDD patients compared to young control participants.

#### Regression analyses

Our findings on confidence ratings and metacognitive efficiency suggest that metacognitive performance is affected primarily by age rather than by the presence of MDD symptoms. To further explore this outcome, we performed a linear regression to predict the mean confidence and metacognitive efficiency of all aged participants (including both aged healthy controls and MDD patients) based on their age, depressive scores, education level and accuracy (accuracy was taken as predictor only for the regression on mean confidence).

A significant regression was found for mean confidence (*F*(4,61) = 3.75, *p* = .009) with an adjusted R^2^ = 0.145. Age (β = -0.02, *p* = .039) and accuracy (β = 0.33, *p* = .037) were significant predictors while education level was marginally significant (β = -0.05, *p* = .064). Depression scores did not predict significantly mean confidence of aged participants (β = - 0.004, *p* = .432). The regression on metacognitive efficiency was marginally significant (*F*(3,62) = 2.365, *p* = .079), with an adjusted R^2^ of 0.06. Education level was the only significant predictor of metacognitive efficiency (β = 0.03, *p* = .041).

## 4. Discussion

An important feature of adaptive behaviour is the ability to judge whether our performance is correct or not, even in the absence of external feedback. This ability is often measured by asking participants to rate the confidence they have in their own performance. Such metacognitive judgements have been suggested to be altered in psychiatric disorders such as MDD (see Hoven et al., 2019 for a review). However, most studies have focused their investigation on the overall under-confidence bias in MDD patients. In contrast, studies investigating the ability for these patients to discriminate correct from incorrect performance (i.e. metacognitive efficiency) are still lacking. In the present study, we used a validated perceptual decision-making task to compare the metacognitive abilities of hospitalized MDD patients and healthy participants while controlling for their objective performance. Because the MDD patients were recruited in an elderly population, and because age has been shown to significantly influence metacognition (Palmer et al., 2014), we also included a third group consisting of healthy young participants to disentangle the effects of age and MDD on metacognitive performance.

Our results showed no difference in overall confidence ratings nor in metacognitive efficiency between MDD patients and their age-matched controls participants. In contrast, we found several indications of a lower metacognitive efficiency in older participants (both healthy aged controls and MDD patients) compared to young participants, suggesting that the ability to discriminate correct from incorrect responses decreases with age and not with depressive symptoms. Furthermore, this discrimination deficit was specifically driven by an overconfidence on incorrect responses.

Our results suggest that MDD is not accompanied with specific deficits of metacognitive efficiency. Two previous studies have reached the same conclusion using the same perceptual task and the same measures of metacognitive performance (Moses-Payne et al., 2019; Rouault et al., 2018). However, these studies investigated metacognition in relation with scores on self-reported questionnaires in general population. In contrast with depressive symptoms, our results showed that the age of our participants was significantly related to their metacognitive efficiency. Specifically, we found that both elderly groups (patients and their aged controls) showed a difficulty to discriminate their performance driven by an overconfidence on errors trials. Note that we initially found overall higher confidence levels in MDD patients and aged controls, suggesting a higher confidence bias in elderly. However, an additional analysis separating confidence for correct and error trials changed our conclusion. The higher confidence in older participants was not systematic of all trials but was specific to errors, indicating a difference of metacognitive efficiency more than confidence bias.

Previous studies have already shown associations between age and overconfidence (Dodson et al., 2007) and between age and lower metacognitive efficiency (Palmer et al., 2014). This finding has important implications as metacognitive deficits can have major consequences for older individuals. Being able to correctly monitor one’s thoughts and performance is essential to behave in a flexible and adaptive manner. This is of particular importance for older individuals as it could help them to compensate for possible age-related deficits in cognitive performance. For instance, an older individual who metacognitively realizes to have some memory difficulties is able to set up remediation strategies (for instance, using a clock or notebook to remind taking in medication).

Surprisingly, we did not find a reduced overall confidence bias in the MDD patients compared to their matched controls. Consistent with the hallmark negative bias in depression (Bradley and Mathews, 1983; McLaughlin and Nolen-Hoeksema, 2011), several non-clinical and clinical studies have found lower confidence levels in depressive patients compared to healthy participants. This tendency for depressive patients to be under-confident has been shown in different domains such as memory, perception or general knowledge (Fieker et al., 2016; Fu et al., 2005; Rouault et al., 2018; Szu-Ting Fu et al., 2012). This negative confidence bias was also shown to correlate with depression severity (Hancock et al., 1996) and improved in recovered patients (Fu et al., 2005). However, a closer look at the existing literature highlights several discrepancies and confounds in previous studies that could explain our contradictory results. First, some studies compared confidence across depressive patients and healthy participants without controlling for task performance (Fu et al., 2005; Szu-Ting Fu et al., 2012). This is problematic because the level of accuracy in the task is intrinsically related to the confidence (Galvin et al., 2003; Pouget et al., 2016; Rouault et al., 2018). When comparing the confidence across groups of individuals, it is therefore crucial to ensure that there is no difference in task performance, as we did with our staircase procedure. A second issue is that many different tasks and measurements have been used to investigate the relations between confidence and depression. Some studies found lower confidence with depression using general knowledge tasks (Stone et al., 2001) or memory recall (Soderstrom et al., 2011) while no effect was found in emotion recognition task (Fieker et al., 2016). One study investigated the relation between confidence and depressive scores across different domains (executive functioning, short-term memory, episodic memory and social cognition) and found no relation between confidence and depression in these tasks, except in the social cognition task (Quiles et al., 2015). The question of the generalizability of metacognitive abilities across cognitive domains is still debated. For instance, metacognition seems to be correlated across different sensory modalities but not between perception and memory (see Rouault, Mcwilliams et al., 2018 for a meta-analysis). It is therefore possible that the influence of depression on confidence depends on the cognitive domain that is investigated. A third issue concerns the heterogeneity of the patients’ sample. While negative affect seems to lower confidence (Culot et al., 2021) the presence of other symptoms such as anxiety could have the opposite effect. Previous studies have found that anxiety has no impact on overall confidence ratings in healthy individuals (Culot et al., submitted; Massoni, 2014; Reyes et al., 2015). Furthermore, overall confidence has been shown to be preserved in a group of both depressed and anxious participants while it was only reduced in “pure” depressed participants (Stone et al., 2001). As many previous studies (Fu et al., 2005; Hancock et al., 1996; Moses-Payne et al., 2019; Rouault et al., 2018; Szu-Ting Fu et al., 2012), we did not assess the presence of anxiety symptoms in our participants, which could have explained the absence of effect on confidence. It is possible that also other sample characteristics such as depression severity, age or medication also partially explain the discrepancies found in the studies on depression and confidence

Besides the absence of anxiety measurement, the present findings should be interpreted in light of other limitations. First, we did not control for the effects of medication, which may have affected metacognitive performance. Previous findings suggest that Lorazepam treatment has no influence on metamemory (Izaute & Bacon, 2005). However, another study hypothesized that second generation of antipsychotic agents might reduce metacognitive biases (Moritz & Woodward, 2004). The effects of antidepressants or anxiolytics on metacognition has never been investigated so far. The size of our patients’ sample does not allow for a comparison of the effects of different pharmacological treatments. This was also not the aim of our study, but deserves investigation in future studies. Second, we did not include some clinical variables in the MDD group such as the number of depressive episodes, illness duration or previous hospitalizations. Third, it is also worth to note that, in the present study, we used a self-reported questionnaire to measure depression severity. However, a potential metacognitive deficit could have biased the insight of the patients into their illness. Despite these limitations, our results point to the importance of age rather than MDD for metacognitive performance. We found no evidence for a specific impact of MDD on confidence bias or metacognitive efficiency. While many studies found a reduced confidence bias with depressive symptoms, the present study suggests that the clinical reality might be more complex. It is possible that the effect of depressive symptoms has been confounded with the effect of other symptoms or sample characteristics. This could have important clinical implications. For instance, two patients with the same traditional MDD diagnosis could show very different metacognitive profiles because they do not have the same anxious symptoms or the same age.

Future clinical studies should therefore aim to investigate the relation between depression (with and without anxiety symptoms) and metacognition in both non-clinical and clinical populations with well validated methodologies and more comprehensive assessments of symptoms.

## Data Availability

All data produced in the present study are available upon reasonable request to the authors

## Author contributions

Catherine Culot was involved in the study design, data collection, data analyses and wrote the first draft of the manuscript. Tina Lauwers, Yamina Madani, Didier Schrijvers and Manuel Morrens contributed to the recruitment and manuscript drafting. Carole Fantini-Hauwel was involved in the study design and manuscript drafting. Wim Gevers contributed to the study design, data analyses and manuscript drafting. All authors read and approved the final version of this manuscript.

## Acknowledgement

The authors thank the University Psychiatric Center Duffel for their help in the data collection. They also thank Netty Pastour and Cassandre Loevenbruck for their help in the data collection of the control participants.

## Footnotes

1 All patients were recruited in a psychogeriatric department. Initially, these patients were recruited as a control group for a study investigating the effects of electroconvulsive therapy in MDD. Electroconvulsive treatment is most often used with elderly MDD patients (Zilles, 2018).

2 Note that a 64-electrodes EEG was also recorded for other purposes and beyond the scope of the current study. A first 6 minutes resting-state was recorded before the task. In addition, EEG was recorded during the metacognitive task.

